# Transformer-based EEG Source Imaging Enables Robust Localization of Pathological High-Frequency Oscillations in Epilepsy

**DOI:** 10.64898/2026.01.14.26344110

**Authors:** Jesse Rong, Zhengxiang Cai, Boney Joseph, Gregory A. Worrell, Bin He

## Abstract

**Objective:** High-frequency oscillations (HFOs) are highly specific biomarkers of epileptogenic tissue, yet their noninvasive localization remains challenging due to their brief duration, low amplitude, and poor signal-to-noise ratio. Here, we introduce TH-DeepSIF, a transformer-based deep learning framework trained on biologically realistic neural mass model simulations, to robustly perform HFO source imaging from scalp EEG.

**Methods:** TH-DeepSIF was evaluated in simulated single- and dual-source HFO scenarios, where it was tasked with recovering both the spatial location and temporal dynamics of HFO generators under increasing spatiotemporal complexity. We further validated TH-DeepSIF in 25 patients with drug-resistant epilepsy by comparing EEG source imaged HFO sources against surgical resection regions.

**Results:** TH-DeepSIF accurately recovered both the spatial location and temporal dynamics of simulated HFO generators, achieving low localization error and strong waveform correspondence with ground truth. TH-DeepSIF localization for pathological HFOs (pHFOs, or spike ripples) demonstrated strong agreement with surgical resection regions, achieving a median localization error of 11.9 mm and specificity of 0.896. Compared with all HFOs (aHFOs), pHFO-based source imaging showed significantly stronger spatial correspondence with resection regions, significantly smaller localization error, and higher precision, sensitivity, geometric mean, and F1 score.

**Significance:** These findings demonstrate that TH-DeepSIF provides a robust, data-driven framework for noninvasive HFO source imaging with improved anatomical specificity and enhanced clinical utility for presurgical evaluation using scalp EEG. Moreover, they show that pathological HFOs (spike ripples )—rather than general HFOs—serve as robust EEG biomarkers for accurate localization of the epileptogenic zone.

**Key Points:**

- A fully data-driven, parameter-free framework for noninvasive HFO source imaging using scalp EEG.
- Pathological HFO source maps exhibit strong spatial concordance with both the surgical resection region and the seizure onset zone.
- Source imaging based on pathological HFOs consistently outperforms imaging based on all detected HFOs.

## Introduction

Epilepsy, a prevalent neurological disorder, affects over 65 million individuals worldwide (1,2). Anti-seizure medications offer relief to approximately two-thirds of the affected population. On the other hand, the removal of the epileptogenic zone (EZ) through a durable clinical procedure offers a pathway to achieving seizure-free status (3–5). This procedure’s success hinges on accurately localizing the EZ before surgery, ensuring minimal tissue removal while attaining seizure freedom. Intracranial EEG (iEEG) yields precise insight into brain regions generating seizure activity (6–8), yet its invasive nature poses surgical risks (9). Furthermore, the intricate process of iEEG electrode insertion prolongs diagnosis and surgical planning, delaying targeted treatment. It is therefore highly desirable to develop non-invasive imaging methods capable of accurately and robustly estimating the EZ to aid surgical planning or optimize iEEG implantation.

Electrophysiological source imaging (ESI) (10,11) enhances the spatial resolution of scalp EEG by estimating the cortical origins of epilepsy biomarkers observed in scalp recordings. Studies demonstrated enhanced spatial resolution of scalp EEG in localizing the EZ from various biomarkers including spikes, seizures and high frequency oscillations (HFOs) (12–18). Regularization techniques are commonly used to solve the underdetermined inverse problem (11). Recent developments have demonstrated the potential of deep learning models trained on synthetic data generated by Neural Mass Models (NMMs) (19,20) to provide precision localization of epileptic sources (21–24). Critically, however, these deep learning approaches have focused almost exclusively on low frequency activities such as spikes or seizures, rather than alternative electrophysiological biomarkers. The emphasis on spikes arises because interictal discharges are shorter, more frequent, and easier to capture within clinical EEG recordings. Yet, it is reported that general spikes have limited spatial specificity and sometimes encounter the challenges arising from multiple spike types (13,25). While seizure localization offers strong capability to localize seizure onset zone (SOZ) (12,14,16,18,24), ictal EEG recordings require extended acquisition time to capture seizures.

High-frequency oscillations (HFOs) (26–29) have become increasingly recognized as clinically important biomarkers for localizing epileptogenic tissue. Studies have suggested that brain regions generating HFOs are more strongly associated with seizure freedom than the irritative zone (IZ) (30). Studies have shown that HFOs co-occurring with interictal spikes—referred to as spike-ripples or pathological HFO (pHFOs)—are more specific to the EZ and better predictors of surgical outcomes than interictal spikes alone (13,31). Despite this growing evidence, clinical use of HFOs in noninvasive analysis remains limited. HFOs are brief, low-amplitude events with poor signal-to-noise ratio (SNR) on scalp EEG, and most established HFO analyses rely on invasive recordings (32,33). Consequently, while interictal spikes are routinely analyzed using ESI, reliable noninvasive source localization of pathological HFOs remains challenging and is not yet widely adopted in clinical practice (10).

Several noninvasive approaches have been proposed to localize HFOs from EEG or MEG, but these methods often require extensive manual tuning, expert-driven component selection, or carefully chosen covariance and thresholding strategies, which may limit scalability across patients and recording conditions (13,34–37). There is therefore a need for a data-driven framework that can directly model the spatiotemporal and spectral characteristics of HFOs from scalp EEG, while reducing dependence on user-defined parameters.

Here, we introduce TH-DeepSIF (Transformer-based High-frequency oscillation Deep Source Imaging Framework) for noninvasive source imaging of HFOs. We evaluate its performance using both controlled simulations and clinical recordings from patients with drug-resistant epilepsy, with localization accuracy assessed relative to clinical ground truth such as surgical resection regions. By learning spatial, temporal, and spectral representations directly from sensor-level data, TH-DeepSIF extends source imaging beyond spikes and seizures, enabling robust imaging cortical origins of pathological HFOs for EZ localization. This parameter-free framework is designed to robustly localize pHFO sources under low SNR and across heterogeneous patient data, enabling scalable and clinically applicable noninvasive HFO source imaging.

## Methods

### TH-DeepSIF Pipeline Overview

We developed TH-DeepSIF which leverages transformer architectures trained on biologically realistic simulations (Fig. 1). The pipeline begins by generating synthetic HFO events using neural mass models (NMM) (19,20) across diverse cortical regions, spatial extents, and waveform morphologies on a 994-brain-region parcellation. These simulated source activities are projected to a 75-channel scalp EEG template using a forward model derived from boundary element modeling (BEM) (38,39). Both the source and scalp signals are high-pass filtered to isolate high-frequency content; the filtered EEG serves as the model input, while the corresponding source activity in ground-truth regions is used as supervision. A transformer model (40) is trained to reconstruct the HFO waveforms and spatial distributions in source space directly from scalp EEG. The trained model is then evaluated on two datasets: a held-out synthetic set with unseen source configurations and real scalp EEG recordings from epilepsy patients with automatically detected HFOs. Performance is assessed against simulated ground truth in synthetic data and clinical ground truth such as surgical resection zones in patient data.

**Figure 1.**
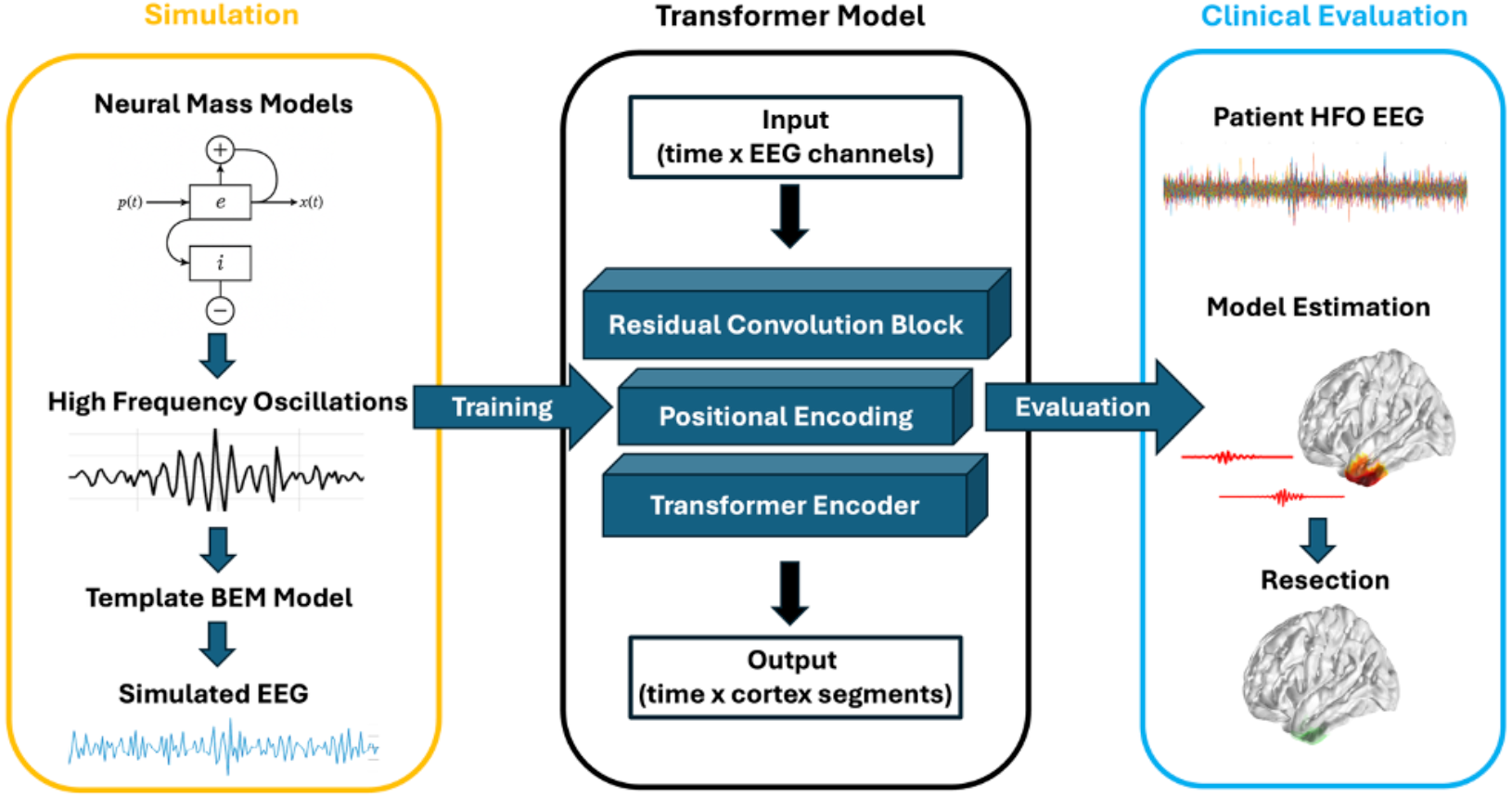
Overview of the TH-DeepSIF pipeline for high-frequency oscillation (HFO) source imaging. (Left) Simulated training data are generated using a neural mass model (NMM), which produces physiologically grounded HFO dynamics at predefined cortical sources. These source-level signals are forward-projected through a realistic head model to simulate corresponding scalp EEG signals. (Middle) A transformer-based deep learning architecture (TH-DeepSIF) is trained end-to-end to learn the mapping from multichannel EEG segments to their underlying cortical source activity. The model leverages self-attention mechanisms to integrate spatial and temporal context across the entire EEG window, enabling robust reconstruction of brief, low-amplitude HFO events. (Right) After training, the network is evaluated on two types of data: (i) held-out simulated HFOs, and (ii) clinically identified HFOs recorded from scalp EEG in patients with epilepsy. This evaluation assesses both generalization to unseen simulations and applicability to real clinical data.

### Simulation of HFOs with Neural Mass Modeling

We developed a physiologically grounded simulation framework based on an extended Jansen–Rit neural mass model (NMM) (19,20,41) to generate HFOs that closely resemble pathological events observed in clinical EEG. The model consists of three interconnected subpopulations—pyramidal neurons, excitatory interneurons, and inhibitory interneurons—whose synaptic gains, time constants, and stochastic input parameters were systematically tuned to produce transient, spatially focal oscillations in the ripple band (>80 Hz). A comprehensive parameter sweep was performed to identify configurations capable of generating realistic waveforms in terms of morphology, dominant frequency, and background activity. These simulated events were evaluated by comparing their spectral and temporal characteristics to those of HFOs recorded with intracranial EEG (Fig. 2).

**Figure 2.**
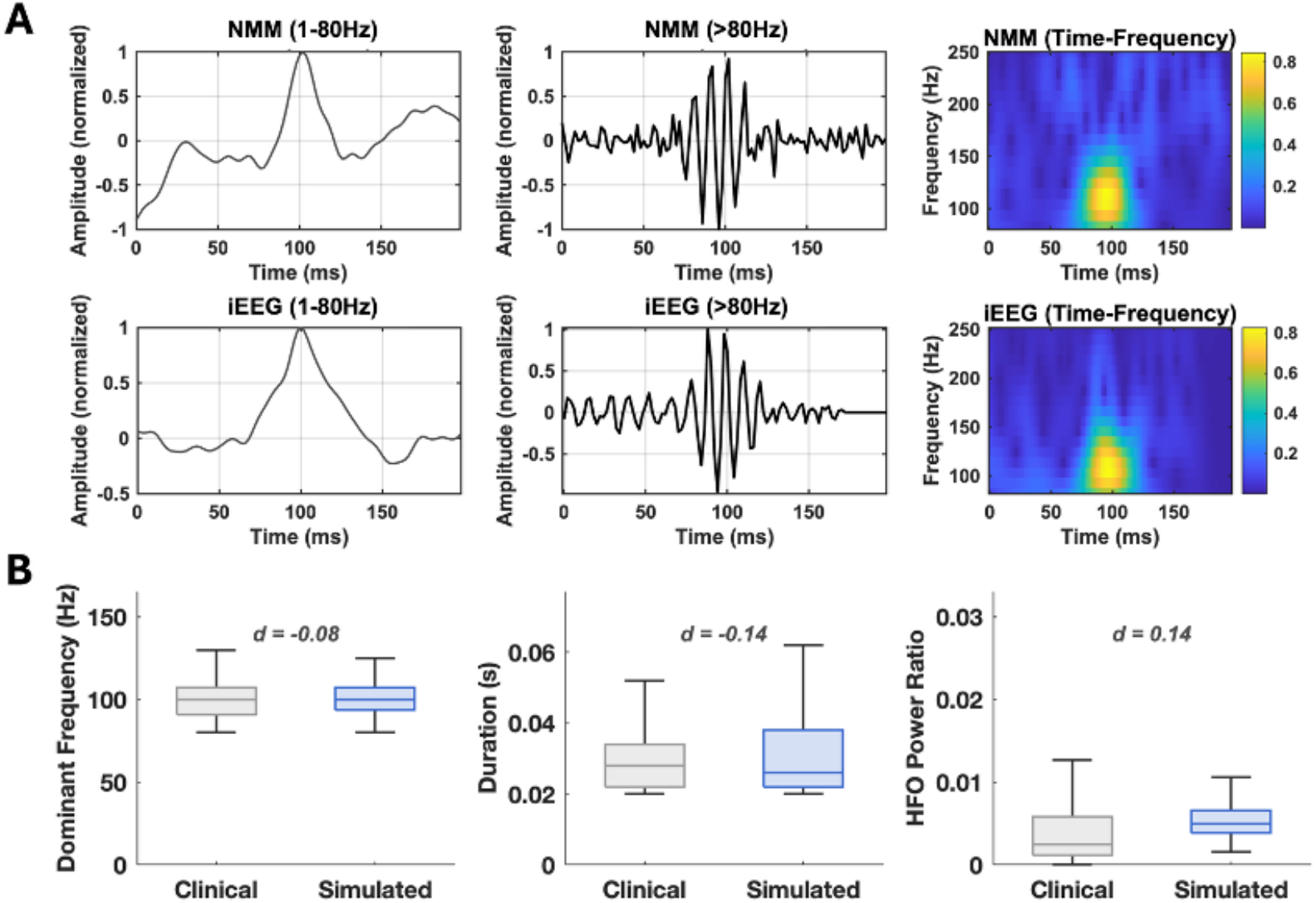
**A**. Comparison between a clinically identified HFO and a simulated HFO generated by the neural mass model (NMM). The top row shows a representative clinical HFO event displayed in three views: wideband EEG (left), high-frequency bandpass (middle), and time-frequency spectrogram (right). The bottom row shows a simulated HFO from the NMM using the same visualization format. **B**. Quantitative comparison of three waveform features between 1,492 clinically identified HFOs from intracranial EEG (iEEG) and 1,116 simulated HFOs generated by the NMM. Features include dominant frequency (left), HFO duration (middle), and the high frequency to broadband power ratio (right). Each panel displays boxplots summarizing the distribution of clinical and simulated HFOs, where the central line indicates the median, the box bounds represent the 25th and 75th percentiles (interquartile range), and the whiskers extend to 1.5 × IQR. Cohen’s d was computed for each feature to quantify the effect size between the clinical and simulated distributions.

HFO sources were embedded within distributed cortical patches defined on the 994-region fsaverage5 template (42), with region connectivity derived from The Virtual Brain framework (43) and spatially extended patches generated using a region-growing procedure (21). Both single-source and dual-source HFO configurations were simulated, with dual-source cases consisting of two spatially distinct cortical regions activated within the same EEG segment. Each one-second EEG segment contained temporally localized HFO activity together with background NMM-derived oscillations. Source activity was forward-projected to the scalp using a 75-channel leadfield matrix generated with a boundary element model implemented in Brainstorm. To emulate realistic clinical conditions, Gaussian white noise was added to the projected EEG at SNRs of 5-20 dB. This pipeline generated a large paired dataset of simulated EEG and ground-truth source activity. In total, 310,128 simulated EEG segments were used for model training, and an independent set of 23,856 simulated segments, generated from NMM parameter configurations not included during training, was reserved for evaluation.

### Transformer-based Deep Neural Network Model Training

We designed a transformer-based deep learning architecture specifically tailored for noninvasive HFO source imaging, accounting for the brief duration and low SNR characteristic of HFO events. The model was trained on a dual-source simulated dataset described above, using EEG segments as input and corresponding source-level time series as targets. Training employed a mean squared error loss function optimized using the Adam algorithm, with model selection based on performance on a held-out validation set. The model was trained on one NVIDIA Tesla V100 GPU at Pittsburgh Supercomputing Center (44).

### Evaluation of Source Reconstruction

To assess generalization, we evaluated the trained network on synthetic datasets containing single and dual HFO sources generated from parameter configurations not included in training. For comparison with a classical distributed source imaging approach, sLORETA was used as a benchmark method (45). The sLORETA inverse solution was computed using a regularization parameter of 0.11, and the resulting inverse filter was applied to the full EEG time series to obtain source-level time series across all cortical regions.

For evaluation, both TH-DeepSIF and sLORETA produced source-level time series for each cortical region. For each detected HFO event, a 200-ms time window centered on the event was extracted from the reconstructed source time series. The magnitude of high-frequency (>80 Hz) source activity within this window was summarized by computing the L2 norm, yielding a single scalar activation value per cortical region, which was subsequently thresholded using Otsu’s method (46) to generate a binary source map for quantitative comparison.

### Clinical Validation

The trained HFO source imaging model was applied to scalp EEG recordings (sampling rate: 500 Hz) from patients with drug-resistant epilepsy. Pathological HFOs (pHFOs), defined as HFOs riding on interictal spikes, were identified using a validated automated detection pipeline previously developed in our laboratory (13). In addition to pHFOs, we defined isolated HFOs as HFO events detected independently of spikes. For analysis, all HFOs (aHFOs) were defined as the union of pHFOs and isolated HFOs. Across the clinical dataset studied, this procedure identified a total of 90 pHFOs and 945 aHFOs.

For each detected HFO, the model generated a cortical source activation map using the same event-centered temporal windowing and norm-based summarization of high-frequency activity described above, which was then registered to the patient’s individual cortical surface reconstructed from pre-operative MRI. All patients had both pre-operative and post-operative MRI, and the post-operative MRI was used to delineate the surgical resection cavity and derive resection regions in the same anatomical space. The spatial relationship between the localized sources and the clinical ground truth, defined by surgical resection regions, was quantified using localization error, precision, specificity, sensitivity, harmonic mean (F1 score) of precision and sensitivity, and the geometric mean of specificity and sensitivity (13,47). iEEG was available in 8 patients, including clinicians defined SOZ.

## Results

### Simulated HFOs Represent Clinical Recordings

To evaluate the biological realism of the simulated HFOs, we compared their temporal and spectral characteristics with representative clinically recorded HFOs. Figure 2A illustrates that the NMM–generated HFOs closely resemble clinical HFOs observed in iEEG across wideband, highpass-filtered, and time–frequency representations. In the wideband signal, both simulated and clinical HFOs exhibited similar oscillatory patterns with major frequency components within the interictal spike range. After high-pass filtering, the simulated HFOs display distinct, transient bursts of narrow-band high-frequency activity (<100 ms in duration) that parallel the morphology and temporal confinement observed in clinical data.

To assess how well the simulated HFOs reproduced the empirical HFO characteristics, we compared the distributions of three waveform features (dominant frequency, event duration, and high-frequency to broadband power ratio) between clinically identified and simulated HFOs (Fig. 2B). Because the clinical and simulated events are independent samples rather than paired measurements of the same events, we focused on distributional similarity rather than formal method-agreement statistics. For each feature, we computed Cohen’s d standardized mean difference between the clinical and simulated datasets (48). Cohen’s d is a widely used effect size that quantifies the magnitude of the difference between two group means in units of the pooled standard deviation. Values of |d| ≈ 0–0.3 are conventionally interpreted as small effects. In our data, all features showed small effect sizes (|d| ≤ 0.15), supporting that the simulated HFOs closely reproduce the empirical feature distributions rather than exhibiting large systematic biases. Figure 2B summarizes group-level quantitative comparisons across three key waveform features: dominant frequency, HFO duration, and high-to-low frequency power ratio. Specifically, dominant frequency yielded an effect size of -0.08, duration −0.14, and high-to-broadband frequency power ratio 0.13, respectively.

Together, these results demonstrated that the simulated HFO dataset faithfully reproduces the core temporal–spectral features of HFOs observed in human epilepsy. Training the deep-learning source imaging model on this biologically grounded dataset is therefore anticipated to promote learning of clinically relevant HFO representations rather than nonspecific high-frequency activity.

### Numerical Evaluation of TH-DeepSIF in Single- and Dual-Source HFO Simulations

Using a model trained on dual-source HFO simulations, we evaluated source imaging performance on two independent synthetic test datasets containing either a single source or two spatially distinct sources, allowing assessment of performance under increasing spatiotemporal complexity. Performance was assessed relative to the known ground truth using sensitivity, precision, localization error (LE), and linear correlation (LC) between reconstructed and ground-truth waveforms (Figure 3). All results reported below are based on synthetic EEG datasets with additive white Gaussian noise applied at the sensor level, with SNRs ranging from 5 to 20 dB, consistent with the noise conditions used during model training.

**Figure 3.**
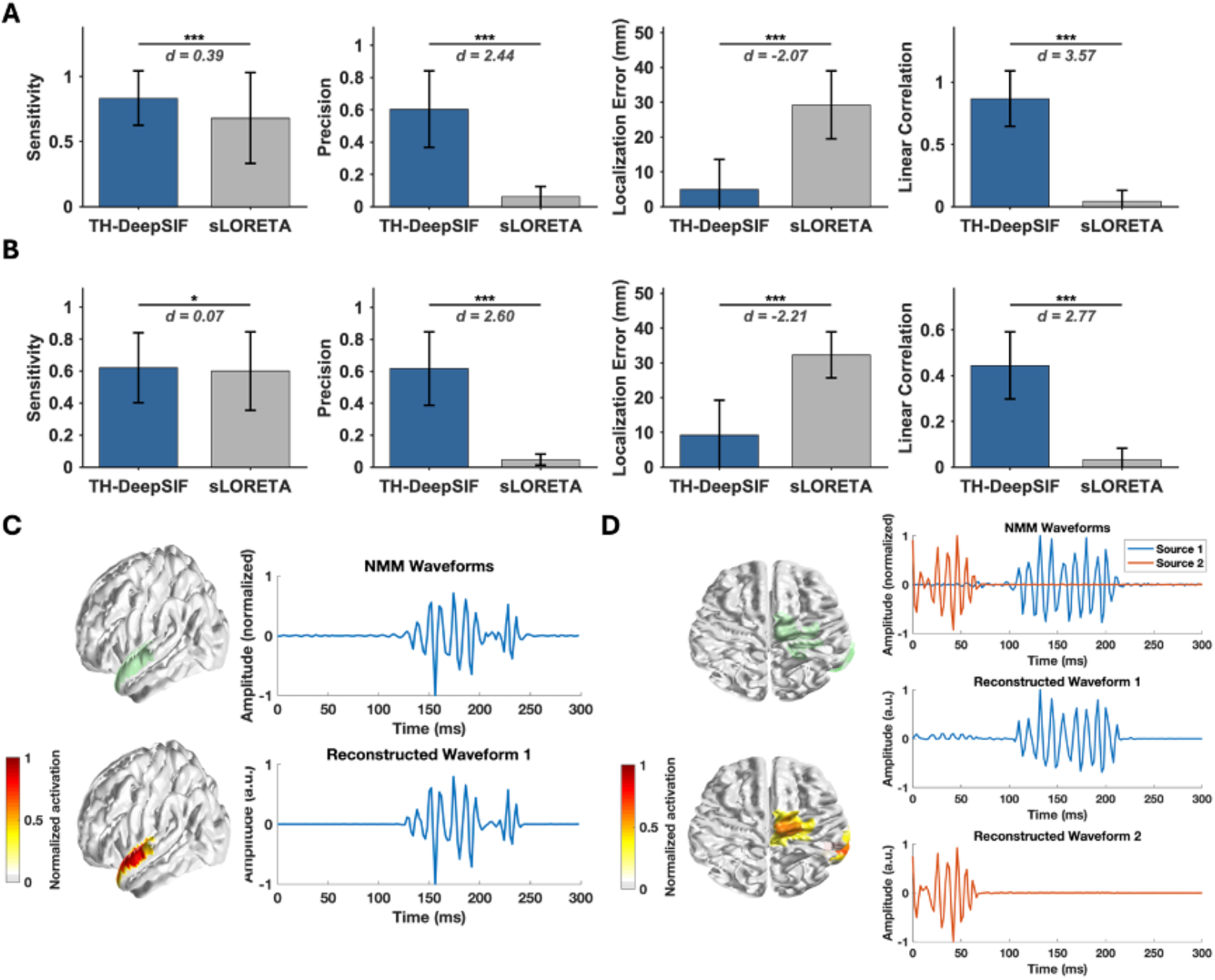
Performances of TH-DeepSIF on simulated HFO datasets, with sLORETA results as comparison. **(A–B)** Group-level metrics (mean ± SD) across 1,000 trials for single-source (A) and two-source (B) conditions. Statistical comparisons between TH-DeepSIF and sLORETA are shown for each metric using Wilcoxon signed-rank tests, with corresponding pairwise p-values and Cohen’s d effect sizes displayed only where applicable; *** p < 0.001, ** p < 0.01. **(C–D)** Representative examples of TH-DeepSIF results in single-source (C) and dual-source (D) scenarios. Left: Example of TH-DeepSIF source imaging result (bottom) vs. ground truth source activity (top, green). Right: Reconstructed vs. ground truth waveforms. TH-DeepSIF accurately localized the spatial generators and reconstructed source-specific waveforms, including disentangling overlapping HFOs in the dual-source case.

In the single-source dataset, TH-DeepSIF achieved high sensitivity (0.839 ± 0.207) and accurately reconstructed the temporal dynamics of HFO activity, yielding a strong waveform correlation with ground truth (Linear Correlation, LC =0.882 ± 0.199). Spatial localization was also strong, with localization error of 4.44 ± 7.29 mm, and precision of 0.632 ± 0.233. TH-DeepSIF reliably recovered both the spatial location and temporal structure of focal HFO sources under single-source conditions.

The dual-source condition introduces increased spatial ambiguity and temporal overlap relative to the single-source case. In the more challenging two-source dataset, overall performance decreased due to increased spatiotemporal overlap; however, TH-DeepSIF continued to reconstruct HFO activity with high temporal fidelity and spatial accuracy. The reconstructed waveforms remained well correlated with ground truth (LC = 0.453 ± 0.136), and spatial estimates remained accurate (LE = 9.36 ± 10.28 mm), with high precision (0.651 ± 0.233). These results demonstrate that TH-DeepSIF can robustly handle multiple concurrent HFO generators while preserving source-specific temporal information.

For reference, we also evaluated sLORETA results using the same datasets and metrics (Figure 3). Across both single- and dual-source conditions, sLORETA showed substantially lower temporal correlation with ground truth, higher localization error, and lower precision than TH-DeepSIF, reflecting more spatially diffuse and temporally imprecise reconstructions.

Figures 3C and 3D provide representative examples of TH-DeepSIF reconstructions under single- and dual-source conditions. In the single-source case (Fig. 3C), the reconstructed source map closely matches the true HFO generator, and the reconstructed waveform aligns with the ground truth in both timing and morphology. In the dual-source case (Fig. 3D), TH-DeepSIF accurately localized both spatially separated sources and successfully disentangled their overlapping temporal activity. Together, these examples illustrate TH-DeepSIF’s ability to jointly model spatial sparsity and temporal dynamics of HFOs, under challenging multi-source conditions.

### TH-DeepSIF Accurately Localizes Pathological HFO Generators in Clinical EEG

We evaluated the clinical relevance of TH-DeepSIF and its performance for EZ localization in a cohort of 25 drug-resistant epilepsy patients, by comparing reconstructed pHFO sources against surgical resection regions using multiple complementary metrics, including localization error, specificity, precision, sensitivity, geometric mean, and F1 score (Figure 4A). These metrics jointly assess localization accuracy, extent estimation accuracy, and the balance between sensitivity and specificity relative to the clinically defined EZ. TH-DeepSIF demonstrated strong source reconstruction performance, with a median localization error of 11.9 mm, indicating close spatial correspondence between reconstructed sources and resection volumes. TH-DeepSIF further achieved high specificity (0.896), reflecting effective suppression of false-positive activations outside the resection region. Precision (0.271) and sensitivity (0.588) values indicate that TH-DeepSIF was able to recover clinically relevant source activity while maintaining reasonable selectivity. Importantly, combined performance measures that account for trade-offs between sensitivity and specificity—such as the geometric mean (of specificity and sensitivity) (0.707) and F1 score (harmonic mean of precision and recall (sensitivity)) (0.679)—were consistently high.

**Figure 4.**
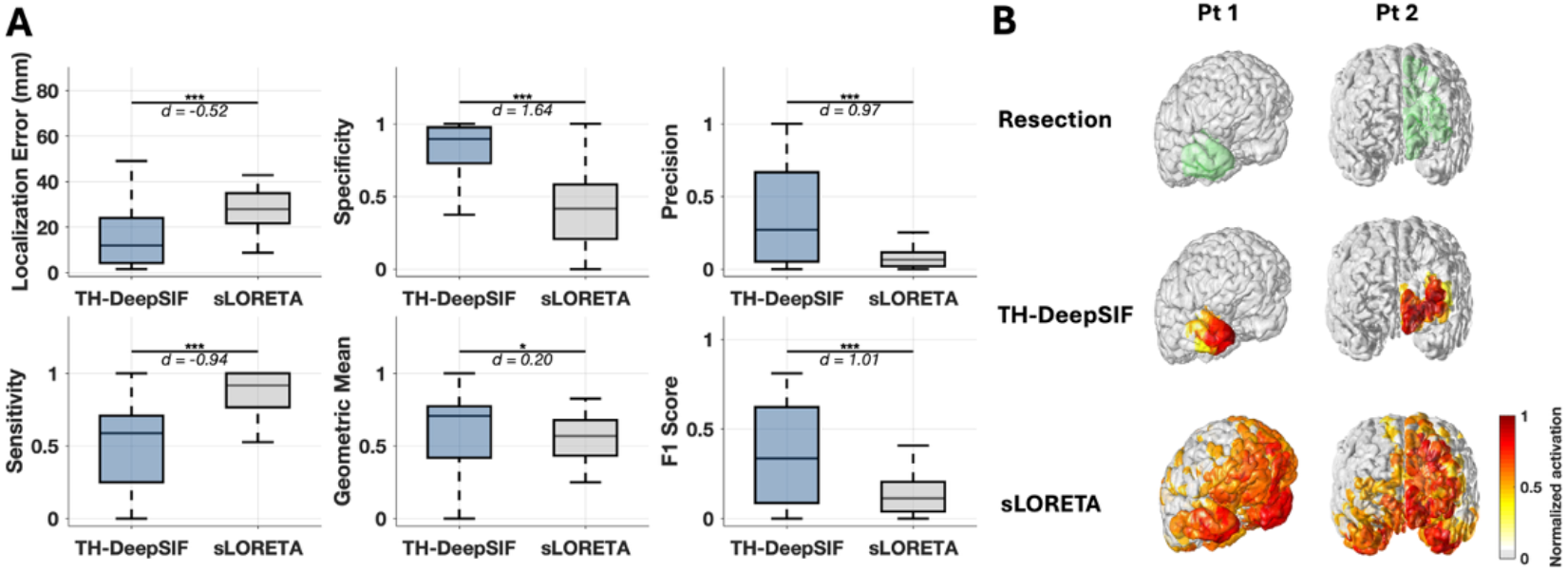
**A**. Quantitative evaluation of imaging performance on 90 pHFO events from 25 patients, comparing TH-DeepSIF and sLORETA. Metrics are computed with respect to the resection regions. Localization error, specificity, precision, sensitivity, geometric mean of specificity and sensitivity and F1 score of precision and recall were computed using Otsu thresholding. Each panel displays boxplots summarizing the distribution of TH-DeepSIF and sLORETA results, where the central line indicates the median, the box bounds represent the 25th and 75th percentiles (interquartile range), and the whiskers extend to 1.5 × IQR. Statistical comparisons were made using the Wilcoxon signed-rank test; *** p < 0.001, ** p < 0.01, * p < 0.05. **B**. Representative source localization results for two HFO events from two patients. The top row shows clinical reference regions: the resected areas (green). The second row displays TH-DeepSIF results and the bottom row presents sLORETA maps. Warmer colors indicate stronger estimated source activity. All source maps were thresholded using the Otsu method.

**Figure 5.**
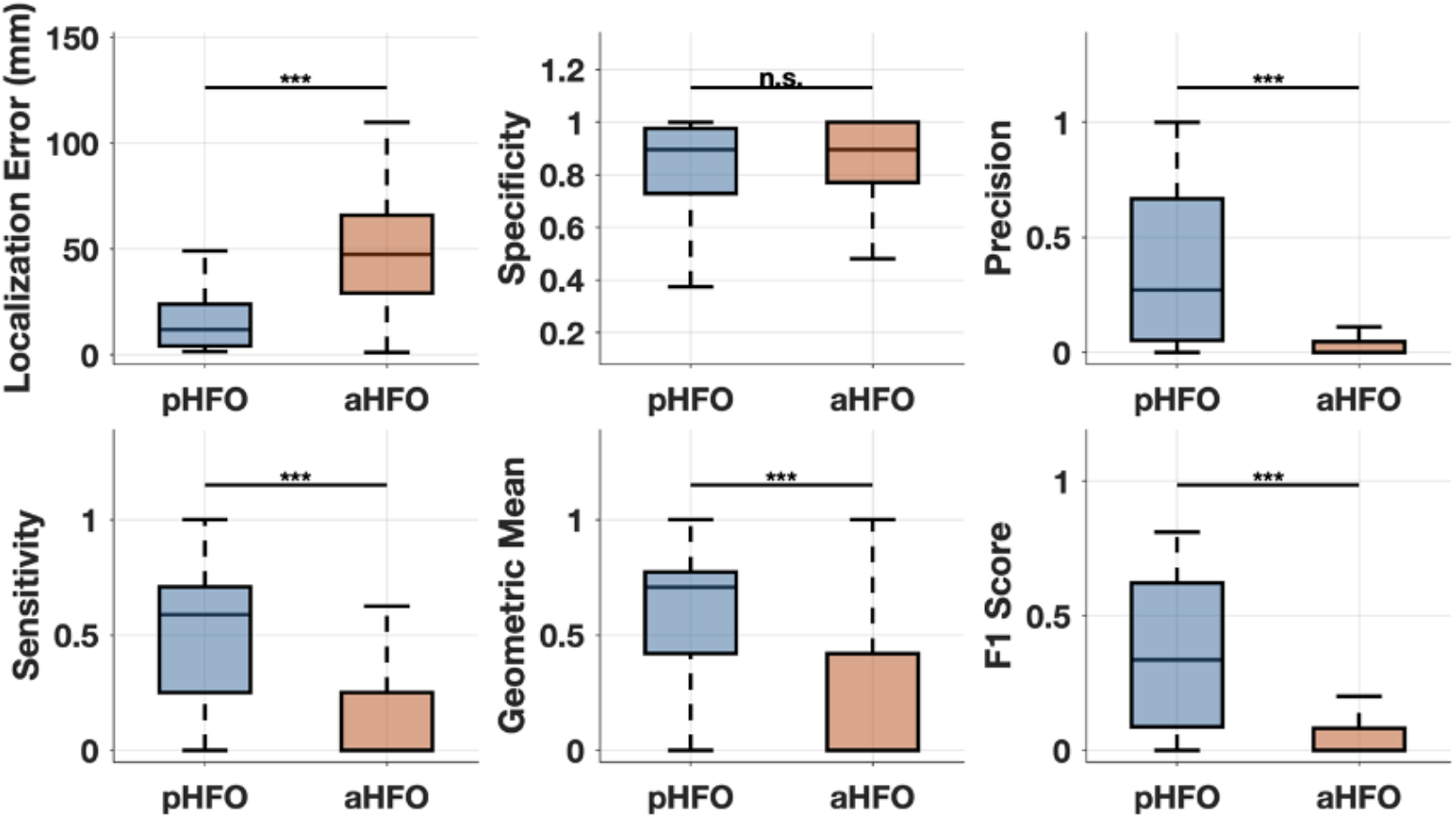
Quantitative comparison of source imaging performance using 90 pathological HFOs (pHFOs) and 945 all detected HFOs (aHFOs) across multiple metrics, including localization error, specificity, precision, sensitivity, geometric mean, and F1 score. Each panel displays boxplots summarizing the distribution of results across events, where the central line indicates the median, the box bounds represent the 25th and 75th percentiles (interquartile range), and the whiskers extend to 1.5 × IQR. Statistical differences between pHFO- and aHFO-based imaging were assessed using the Wilcoxon rank-sum test. *** p < 0.001; “n.s.” p > 0.05.

We further compared TH-DeepSIF with sLORETA, a classical distributed source imaging method serving as a well-established benchmark (Figure 4A). TH-DeepSIF achieved significantly lower localization error than sLORETA (11.9 vs 27.9 mm, p < 0.001), indicating improved spatial accuracy. TH-DeepSIF also showed significantly higher specificity (0.896 vs 0.417, p < 0.001) and precision (0.271 vs 0.065, p < 0.001), indicating that its reconstructed sources were more spatially confined to the resection region with substantially fewer false-positive activations outside clinically relevant cortex, whereas sLORETA produced more spatially diffuse estimates that extended beyond the epileptogenic zone. Although sLORETA demonstrated higher sensitivity (0.917 vs 0.588, p < 0.001), this was accompanied by substantially lower specificity and increased localization error, consistent with more spatially diffuse source reconstructions. Overall, TH-DeepSIF achieved significantly higher geometric mean (0.707 vs 0.568, p = 0.013) and F1 score (0.336 vs 0.113, p < 0.001), indicating improved balance between sensitivity and specificity as compared to sLORETA.

Examples from two representative pHFO events (Figure 4B) further illustrate these differences. TH-DeepSIF consistently localized HFO-associated activity within or immediately adjacent to the surgical resection zone, generating compact and anatomically plausible source maps. In contrast, sLORETA produced spatially diffuse activation patterns that frequently extended beyond regions of clinical relevance, reducing interpretability and target specificity.

Together, these findings demonstrate that TH-DeepSIF enables accurate, spatially focal, and temporally coherent noninvasive localization of pathological HFO generators. By producing compact source estimates that align closely with surgical resection regions and preserve the expected spike–HFO temporal coupling, TH-DeepSIF provides clinically meaningful source reconstructions that are more anatomically specific than conventional distributed methods, supporting its relevance for HFO-based EZ localization.

### Comparison Between Pathological HFOs (pHFOs) and All Detected HFOs (aHFOs)

To examine how the physiological type of HFO influences noninvasive source localization, we compared source imaging results obtained using pathological HFOs coupled to interictal spikes (pHFOs) and all detected HFOs (aHFOs). pHFOs were defined as HFOs temporally overlapping with interictal spikes, whereas aHFOs included both spike-coupled and isolated HFOs occurring independently of spikes. Across the clinical dataset examined, a total of 90 pHFOs and 945 aHFOs were identified. Because the proposed framework is designed to localize HFO-related activity broadly, this analysis evaluated whether source localization accuracy and overlap with surgical resection differ when using a more selective pHFO subset versus the full set of detected HFO events.

Consistent with this expectation, pHFO-based source imaging showed significantly stronger spatial correspondence with resection regions than aHFO-based imaging across multiple metrics (Wilcoxon rank-sum tests, p < 0.001 for localization error, precision, sensitivity, geometric mean, and F1 score). Localization error was substantially lower for pHFOs than for aHFOs (median [IQR]: 11.9 [4.2–24.0] mm vs. 47.4 [29.2–65.9] mm), indicating greater spatial accuracy when restricting analysis to spike-coupled pHFO events.

Differences were particularly pronounced in metrics for extent estimation. pHFO-based imaging yielded significantly higher precision (median [IQR]: 0.271 [0.053–0.667]) and sensitivity (0.588 [0.250–0.708]) compared with aHFO-based imaging (precision: 0.000 [0.000–0.046]; sensitivity: 0.000 [0.000–0.250]). As a result, combined performance measures that balance sensitivity and specificity were also significantly higher for pHFOs, including the geometric mean (0.707 [0.419–0.773] vs. 0.000 [0.000–0.419]) and F1 score (0.336 [0.087–0.622] vs. 0.000 [0.000–0.082]).

Importantly, both pHFO- and aHFO-based source estimates were generally focal rather than spatially diffuse, as reflected by uniformly high specificity values in both groups despite the small size of surgical resection regions relative to the cortical surface. This indicates that neither approach produced widespread spurious activation, and that false-positive suppression was comparable between pHFOs and aHFOs. Although specificity differed slightly between groups (median [IQR]: 0.896 [0.729–0.976] for pHFOs vs. 0.896 [0.771–1.000] for aHFOs; Wilcoxon rank-sum p = 0.092, the small magnitude of this difference and the substantial overlap of distributions suggest minimal practical impact. Instead, the primary distinction between the two conditions lay in where these focal sources were localized. While pHFO-based imaging preferentially localized to regions within or near the resection, aHFO-based imaging frequently localized to focal cortical areas outside the resection region. This pattern is consistent with the heterogeneous physiological origins of aHFOs, which may arise from non-epileptogenic or spatially distinct generators that remain focal but are not part of the surgically targeted epileptogenic zone.

Overall, these results indicate that restricting analysis to spike-coupled pathological HFOs yields more consistent and spatially accurate noninvasive source localization, reflecting the greater focality of their underlying generators. In contrast, the heterogeneous origins of aHFOs result in increased variability in source estimates, which limits their reliability for resection-level correspondence when analyzed in aggregate.

## Data Availability

All data produced in the present study are available upon reasonable request to the authors

## Discussion

Accurate noninvasive localization of pathological high-frequency oscillations (pHFOs) holds substantial promise for improving presurgical evaluation in drug-resistant focal epilepsy by providing biomarkers with high specificity and low localization error for the epileptogenic zone (EZ) (26,27,30,33,34). In this study, we demonstrate that TH-DeepSIF can localize pHFO-related activity from scalp EEG with strong spatial accuracy and temporal precision. Localization performance was quantitatively validated against known ground truth in simulations and assessed in patients through concordance with surgical resection regions (15,16,32). Across simulations and clinical data, TH-DeepSIF consistently produced focal source estimates with low localization error and high specificity, supporting its effectiveness for noninvasive HFO source imaging.

This work specifically focuses on pHFOs—high-frequency oscillations temporally coupled to interictal spikes—which have been shown to outperform spikes or HFOs alone in identifying epileptogenic tissue (13,25,31,47,49). Consistent with this literature, we observed substantially improved localization performance for pHFO-based imaging compared with imaging based on HFOs alone (aHFOs). In clinical data, pHFO reconstructions exhibited markedly lower localization error and higher precision and specificity, whereas aHFO-based reconstructions frequently failed to produce spatially meaningful overlap with resection regions. The poor performance of aHFOs likely reflects their heterogeneous origins and lower pathological specificity, as high-frequency activity not temporally coupled to spikes may arise from physiological or nonspecific sources (30,33,50). These findings reinforce the importance of incorporating spike–HFO coupling to improve the reliability and interpretability of noninvasive HFO source imaging.

Previous noninvasive HFO imaging studies have shown that advanced methodological pipelines can achieve strong performance when supported by expert-driven processing. Tamilia et al., for example, used beamforming and ripple-onset analyses with high-density EEG/MEG to localize epileptogenic regions and improve outcome prediction (36). More recently, Jiang et al. proposed a spatial–temporal–spectral source imaging (STSI) framework that jointly leverages spatial, temporal, and spectral features of EEG biomarkers—including HFOs and spike–HFO events—to improve localization accuracy relative to clinical references such as surgical resection and SOZ (47). In contrast, TH-DeepSIF is fully parameter-free: it operates directly on a single high-pass–filtered EEG segment and automatically learns the spatial and temporal representations needed for HFO and spike-ripple localization, without requiring manual selection of covariance windows, regularization strategies, or spatial constraints. This design enables data-driven extraction of waveform morphology, temporal context, and spatial sparsity within a unified architecture, reducing dependence on user expertise and enhancing scalability across diverse HFO morphologies.

Several limitations should be noted. Surgical resection provides only an indirect approximation of the true EZ, and clinical outcomes may be influenced by factors beyond HFO localization (1). To address this, we evaluated multiple complementary metrics and validated performance across both simulated and clinical datasets. In addition, the modest precision observed in clinical evaluations reflects the intrinsically low signal-to-noise ratio of scalp-recorded HFOs, a well-recognized limitation of noninvasive HFO imaging that has been consistently reported in prior studies (28,34,37). Lastly, although TH-DeepSIF itself is parameter-free, the overall pipeline relies on preprocessing steps for event detection, which may introduce variability. Future work integrating end-to-end learning from longer EEG segments or coupling TH-DeepSIF with automated detection frameworks may further improve robustness and scalability.

## Author Contributions

Conceptualization: J.R. and B.H.; Methodology: J.R., Z.C. and B.H; Data curation: GAW, B.J.; Formal analysis and visualization: J.R.; Investigation: J.R., Z.C., and B.H.; Writing—original draft: J.R.; Writing—review & editing, J.R., Z.C., and B.H.; Supervision: B.H.

## Acknowledgments

This work was supported in part by NIH grants NS096761, NS127849, NS131069, NS124564, and EB029365. The authors are grateful to Dr. Xiyuan Jiang and Colton Gonsisko for useful discussions.

